# Measuring Frailty Using Self-Report or Automated Tools to Identify Risk of Cardiovascular Events and Mortality: The Million Veteran Program

**DOI:** 10.1101/2023.10.18.23297231

**Authors:** Saadia Qazi, Benjamin Seligman, Sarah R. Preis, Manas Rane, Luc Djousse, David R. Gagnon, Peter W.F. Wilson, J. Michael Gaziano, Jane A. Driver, Kelly Cho, Ariela R. Orkaby, MVP

**Author notes:** Corresponding Author: Saadia Qazi DO, MPH, Divisions of Cardiovascular Disease and Aging Brigham and Women’s Hospital, Boston VA Medical Center, 150 S Huntington Avenue Boston, MA 02130.

## Abstract

**Background:** Frailty, a syndrome of physiologic vulnerability, increases cardiovascular disease (CVD) risk. Whether in person or automated frailty tools are ideal for identifying CVD risk remains unclear. We calculated three distinct frailty scores and examined their associations with mortality and CVD events in the Million Veteran Program (MVP).

**Methods:** MVP is a prospective cohort of U.S. Veterans that has enrolled nearly one million Veterans. We included participants aged ≥50 years who enrolled from 2011-2018. Frailty was calculated using three tools: two frailty indices (FI) based on the accumulation of deficits theory, the 36-item MVP-FI using self-reported answers to questionaries, and the 31-item VA-FI developed using claims data. Finally, we calculated the 3-item Study of Osteoporotic Fractures Fried physical frailty score from self-report. The primary outcomes were CVD and all-cause mortality. Multivariable-adjusted Cox proportional hazards models (adjusted for age, sex, smoking, statin use, antihypertensive use, hyperlipidemia, and survey year). Secondary outcomes were myocardial infarction (MI), stroke, and heart failure (HF).

**Results:** Among 190,688 MVP participants (mean age 69 ±9 years, 94% male, 85% white), there were 33,233 (17%) all-cause and 10,115 (5%) CVD deaths. Using MVP-FI, 29% were robust, 42% pre-frail, and 29% frail. Frailty prevalence increased by age group, from 27% in 50–59-year-olds to 42% in age≥90 years. Follow-up duration was 6±2 years. Using the MVP-FI, pre-frail and frail Veterans had a higher incidence of both all-cause mortality (pre-frail: HR=1.66, 95%CI: 1.61-1.72; frail: 3.05, 2.95-3.16) and CVD death (pre-frail: 1.76, 1.65-1.88; frail: 3.65, 3.43-3.90), as compared to robust individuals. These findings remained significant among Veterans ≥ 50 years. Findings were similar for CVD events. When frailty was measured using the VA-FI and SOF results were concordant.

**Conclusion:** Irrespective of frailty measure used, frailty is associated with a higher risk of all-cause mortality and CVD events. Further study of frailty in individuals <60 years old is warranted.

## INTRODUCTION

Cardiovascular disease (CVD) is the leading cause of death among older adults in the United States, responsible for over 750,000 deaths among adults aged ≥65 in 2020.^1^ Managing this burden is critical to meet *Healthy People* 2030 goals for coronary heart disease and stroke deaths.^2^ However, major risk calculators, such as the Framingham Risk Score, Reynold’s Risk Calculator, and QRISK-3 have maximum ages of 79, 80, and 84 respectively, while the Pooled Cohort Equations have a maximum age of 75. Risk-stratification of heart disease risk among older adults is challenging, in part due to physiologic aging that includes inflammation, altered metabolism, genomic alterations, and other changes not included in traditional risk calculators, as well as competing risks from other causes of death.^3–5^

Frailty may address the challenges of both aging physiology and competing risk when assessing CVD risk in older adults. Defined as the vulnerability to poor health outcomes following a physiologic stressor, frailty is operationalized according to two leading theories:^6^ 1) physical frailty, which treats frailty as a discrete syndrome of weight loss, exhaustion, fatigue, decreased physical activity, and weakness;^7^ and 2) deficit accumulation frailty which considers health-related deficits spanning morbidity, cognition, function, nutrition and mental health.^8^ Both approaches have been associated with major adverse cardiovascular outcomes (MACE) across the spectrum of CVD and may inform prognostication and management of patients.^9–12^ The application of frailty in the management of CVD patients first became standard in aiding risk stratification for transcatheter aortic valve replacement (TAVR) procedures and has subsequently expanded to heart failure prognostication and management and other areas of CV care.^13,14^ Despite the recognition of frailty as a critical tool for prognostication and management in CVD, among the 60 frailty tools developed based on these two theories of frailty, multi-society guideline recommendations lack specific guidance on which available tool is best.^15,16^ In an effort to address the question of optimal frailty assessment we used the Million Veteran Program (MVP), a large, well-phenotyped cohort of over 900,000 US Veterans. We used both a deficit accumulation and a physical frailty approach to examine the relationship between frailty, mortality, and CVD events.

## METHODS

### Study Sample

The Million Veteran Program (MVP) is a prospective observational cohort study of US Veterans, which was established to explore the effects of genetics, lifestyle, and military exposures on veteran health.^17^ Starting in 2011, veterans who received healthcare through the Veterans Health Association (VHA) were invited to enroll in MVP. Participants completed the MVP Baseline and Lifestyle surveys, provided a blood sample, and allowed access to their VHA health records through the VA’s Corporate Data Warehouse (CDW) repository.^18^ This study was approved by the VA Central Office Institutional Review Board (IRB) in Washington, DC. A waiver for patient consent was obtained from the VA Boston Healthcare system IRB.

For the present study, we included all participants aged ≥50 years who completed both the Baseline and Lifestyle Surveys, during the period of 2011-2018. Participants were excluded if they completed their MVP Lifestyle Survey more than 30 days before or one year after their MVP Baseline Survey, were missing more than 20% of the necessary information to calculate the MVP frailty score, covariates, or date of cardiac event, or died prior to completion of the questionnaire (Supplemental Figure 1).

### Frailty Assessments

#### MVP Frailty Index (MVP-FI)

We used the MVP Baseline and Lifestyle Surveys to develop a deficit accumulation frailty index, the “Million Veteran Program Frailty Index” (MVP-FI), based on a standard procedure using self-reported answers to the questionnaires.^19^ This index is composed of 33 deficits covering health status and function, mental health and cognition, and comorbidities. The specific deficits and the point values assigned to the survey responses are listed in Supplemental Table 1. To calculate the MVP-FI, the number of deficits present for an individual were summed and divided by the number of deficits assessed, producing a frailty index value between 0 and 1. If a participant had six or fewer missing responses to the survey items, their MVP-FI was calculated as the number of deficits present divided by the total number of items completed. Individuals with seven or more missing items were excluded. We defined an MVP-FI of 0 to 0.10 as robust, 0.11 to 0.20 as pre-frail, and >0.20 as frail, consistent with prior frailty measures developed within the VA such as the Veterans Affairs Frailty Index (VA-FI).^20^

#### Other Frailty Measures

To compare the MVP-FI, which is based solely on self-reported measures, with an automated deficit accumulation frailty measure previously developed using VA medical records, we also calculated the VA-FI.^11,20,21^ The VA-FI uses International Classification of Diseases (ICD) and the Healthcare Common Procedure Coding System (HCPCS) codes from the VA electronic medical records and Medicare claims data to assess 31 deficits including physical health, mental health and cognition, and function. It has been extensively validated in the VA population and associated with all-cause and cardiovascular mortality.^11,20^ We calculated the VA-FI using clinical claims data from the 36 months preceding completion of the MVP Baseline Survey.

To compare a deficit accumulation frailty index with a physical frailty measure, we operationalized the Study of Osteoporotic Fractures frailty index (SOF) using questionnaire items from the MVP Baseline and Lifestyle Surveys. The original SOF is composed of three items scored as 0 or 1: weight loss, inability to rise from a chair five times without using arms, and reduced energy.^22^ The MVP surveys contained specific questionnaire items pertaining to weight loss and reduced energy, however, the number of chair stands was not assessed. As a proxy we used the participants’ responses to the ADL question “Required assistance with transferring from a bed or chair in the last week.” This is similar to other modified versions of the SOF.^23^ The specific survey questions and coding are presented in Supplemental Table 2. The SOF scores ranged from 0 to 3. A participant was considered robust if they had a score of 0, pre-frail if they had a score of 1, and frail if they had a score of 2 or 3.

### Outcomes

The primary outcomes were CVD and all-cause mortality. Deaths were identified through the National Death Index (NDI). A death was classified as CVD-related if one of the following ICD codes was listed as the primary cause of death in the NDI (ICD-10 codes: I10-I16, I20-I25, I27-I28, I34-I37, I44-I51, I60-I75, I77-I78, I42, I97, I99, R58, G45, R00). Secondarily we considered the outcomes of incident ischemic stroke (ICD-9 codes: 433.x1, 434 (excluding 434.x0), 436, 437.0, 437.6; ICD-10 codes: I63.xx9, I63.20, I63.22, I63.50, I63.59, I67.2, I67.6, I67.89), myocardial infarction (MI; ICD-9 codes: 410.x, 411.x, 412.x, 414.x; ICD-10 codes: I20.0, I21.x, I22.x, I24.x, I25.x), and heart failure (ICD-9 code: 428.x; ICD-10 code: I50). ^24^ Secondary outcomes were derived from the VA electronic medical record data linked with Medicare and Medicaid claims.

### Clinical Covariates

Information on age, sex, race, and ethnicity was extracted from the MVP Baseline Survey. Smoking status was derived from the MVP Baseline Survey and VHA medical records using a previously published algorithm.^25^ History of hyperlipidemia (ICD-9 code 272.4; ICD-10 codes: E74.8, E78.5), antihypertensive medication (VA National Formulary classification: CV100, CV150, CV200, CV400, CV490, CV701, CV702, CV704, CV800, CV805, CV806), and statin use were determined using VHA medical records. Medication use was assessed during the 24 months prior to the date of a participant’s MVP Baseline Survey date.

### Statistical Analysis

Descriptive statistics were calculated using means (standard deviation), medians (interquartile range), or frequency counts and percentages, as appropriate. Spearman correlation coefficients were calculated to examine the correlation between the MVP-FI, VA-FI, and SOF frailty measures. Cox proportional hazards models were used to obtain hazards ratios (HR) and 95% confidence intervals (CI) for the association between each frailty measure and the outcomes. All models were adjusted for age, sex, smoking status, history of hyperlipidemia, antihypertensive treatment, statin use, number of days between the return date of the MVP Baseline Survey and Lifestyle Survey, and the MVP Baseline Survey calendar year. Because the secondary outcomes comprised incidence of CVD events, participants with MI were excluded from models assessing incidence of MI. Similar exclusions were applied for models investigating incidence of stroke and HF. The analysis was conducted using SAS Enterprise v. 7.1. A p-value of <0.05 was considered statistically significant.

## RESULTS

The total study population included 190,688 Veterans as shown in Supplemental Figure 1. Mean age of participants in the overall study was 69 ±9 years, 11,852 (6%) were female, 161,900 (85%) identified as white and 16,344 (9%) as Black. At baseline, a history of stroke was present in 5% (n=9,397), MI in 7% (n=13,254) and HF was present in 9% (n=17,714) of Veterans. Hyperlipidemia, treatment of hypertension, statin treatment, history of CVD, and history of HF were more common at higher levels of frailty (Table 1).

**Table 1.** Baseline characteristics and outcomes of 190,688 participants of the Million Veteran Program.

|  | MVP-FI Frailty Status |  |  | Total<br>(190,688) |
| --- | --- | --- | --- | --- |
|  | Robust<br>(N=55,487) | Pre-Frail<br>(N=79,835) | Frail<br>(N=55,366) |  |
| MVP Frailty Index Score |  |  |  |  |
| Mean ± SD | 0.064±0.026 | 0.149±0.028 | 0.282±0.078 | 0.163±0.098 |
| Median (25<sup>th</sup>, 75<sup>th</sup> percentile) | 0.068 (0.045, 0.083) | 0.148 (0.121, 0.174) | 0.258 (0.227, 0.313) | 0.148 (0.091, 0.212) |
| Minimum, maximum | 0.000, 0.100 | 0.102, 0.200 | 0.202, 0.914 | 0.000, 0.914 |
| SOF Index |  |  |  |  |
| Robust (0) | 39338 (72.12) | 43545 (55.59) | 15551 (28.89) | 98434 (52.73) |
| Pre-frail (1) | 14354 (26.32) | 29467 (37.62) | 26346 (48.95) | 70167 (37.58) |
| Frail (2,3) | 854 (1.57) | 5314 (6.78) | 11923 (22.15) | 18091 (9.69) |
| VA-FI |  |  |  |  |
| Robust | 41051 (73.98) | 30107 (37.71) | 6846 (12.36) | 78004 (40.91) |
| Pre-frail | 11790 (21.25) | 33626 (42.12) | 19212 (34.7) | 64628 (33.89) |
| Frail | 2646 (4.77) | 16102 (20.17) | 29308 (52.94) | 48056 (25.2) |
| Years of Follow-up, Mean ± SD | 6.4±2.0 | 6.2±2.2 | 5.7±2.4 | 6.1±2.2 |
| Age (years), Mean ± SD | 68±9 | 70±9 | 70±9 | 69±9 |
| Age category, n (%) |  |  |  |  |
| 50-59 years | 9302 (16.76) | 10042 (12.58) | 7304 (13.19) | 26648 (13.97) |
| 60-69 years | 24276 (43.75) | 33105 (41.47) | 23086 (41.7) | 80467 (42.2) |
| 70-79 years | 15974 (28.79) | 23958 (30.01) | 15463 (27.93) | 55395 (29.05) |
| 80-89 years | 5372 (9.68) | 11143 (13.96) | 7981 (14.41) | 24496 (12.85) |
| ≥90 years | 563 (1.01) | 1587 (1.99) | 1532 (2.77) | 3682 (1.93) |
| Female sex, n (%) | 3330 (6.00) | 4725 (5.92) | 3797 (6.86) | 11852 (6.22) |
| Race, n (%) |  |  |  |  |
| White | 48552 (87.5) | 68341 (85.6) | 45007 (81.29) | 161900 (84.9) |
| Black/African-American | 4039 (7.28) | 6752 (8.46) | 5543 (10.01) | 16334 (8.57) |
| Asian | 405 (0.73) | 486 (0.61) | 336 (0.61) | 1227 (0.64) |
| Native Hawaiian/Pacific Islander | 53 (0.10) | 97 (0.12) | 87 (0.16) | 237 (0.12) |
| American Indian/Alaska Native | 203 (0.37) | 441 (0.55) | 524 (0.95) | 1168 (0.61) |
| Hispanic/Latino ethnicity | 2811 (5.08) | 4081 (5.13) | 3545 (6.42) | 10437 (5.49) |
| Multi-race | 1405 (2.53) | 2482 (3.11) | 2643 (4.77) | 6530 (3.42) |
| Other | 725 (1.31) | 1102 (1.38) | 1128 (2.04) | 2955 (1.55) |
| Unknown | 105 (0.19) | 134 (0.17) | 98 (0.18) | 337 (0.18) |
| Smoking status, n (%) |  |  |  |  |
| Never smoker | 19705 (35.51) | 21931 (27.47) | 12606 (22.77) | 54242 (28.45) |
| Former smoker | 27384 (49.35) | 43183 (54.09) | 30234 (54.61) | 100801 (52.86) |
| Current smoker | 8398 (15.14) | 14721 (18.44) | 12526 (22.62) | 35645 (18.69) |
| Hypercholesterolemia, n (%) | 34424 (62.04) | 57808 (72.41) | 44203 (79.84) | 136435 (71.55) |
| Statin treatment*, n (%) | 21247 (38.29) | 42432 (53.15) | 36534 (65.99) | 100213 (52.55) |
| Antihypertensive treatment*, n (%) | 24917 (44.91) | 53442 (66.94) | 44789 (80.9) | 123148 (64.58) |
| Baseline survey year, n (%) |  |  |  |  |
| 2011-2012 | 11869 (21.39) | 18291 (22.91) | 12867 (23.22) | 43027 (22.56) |
| 2013-2014 | 18516 (33.37) | 26165 (32.77) | 18014 (32.51) | 62695 (32.87) |
| 2015-2016 | 14370 (25.9) | 20690 (25.91) | 14821 (26.75) | 49881 (26.15) |
| 2017-2018 | 10735 (19.35) | 14702 (18.41) | 9703 (17.51) | 35140 (18.42) |
| History of stroke at baseline, n (%) | 767 (1.38) | 3118 (3.91) | 5512 (9.96) | 9397 (4.93) |
| History of MI at baseline, n (%) | 1104 (1.99) | 4653 (5.83) | 7497 (13.54) | 13254 (6.95) |
| History of CHF at baseline, n (%) | 1269 (2.29) | 5674 (7.11) | 10771 (19.45) | 17714 (9.29) |

| *Outcomes* |  |  |  |  |
| --- | --- | --- | --- | --- |
| All-cause mortality, n (%) | 4717 (8.50) | 13060 (16.36) | 15446 (27.9) | 33223 (17.42) |
| Cardiovascular mortality, n (%) | 1217 (2.19) | 3775 (4.73) | 5123 (9.25) | 10115 (5.3) |
| Incident stroke, n (%) | 405 (0.74) | 997 (1.30) | 998 (2.00) | 2400 (1.32) |
| Incident MI, n (%) | 531 (0.98) | 1482 (1.97) | 1580 (3.30) | 3593 (2.02) |
| Incident CHF, n (%) | 2344 (4.32) | 6587 (8.88) | 6885 (15.44) | 15816 (9.14) |

Abbreviations: SOF, Study of Osteoporotic Fractures FI, frailty index; MI, myocardial infarction; CHF, congestive heart failure.

Overall, 55,487 (29%) of the sample was robust by the MVP-FI, 79,835 (42%) were pre-frail, and 55,366 (29%) were frail. The distribution by VA-FI was similar, with 78,004 (53%) robust, 64,628 (34%) pre-frail, and 48,056 (25%) frail. Using physical frailty as assessed by SOF, 98,434 (53%) were robust, 70,167 (38%) were pre-frail, and 18,091 (10%) were frail. While 86% of included participants were aged ≥ 60 years, 26,248 (14%) were aged 50-59 years, of which 13% were pre-frail and another 13% frail (Table 1). Spearman correlation coefficients between the three frailty indices were as follows: continuous FI scores for MVP FI and SOF = 0.39 (p<0.0001), MVP FI and VAFI = 0.61 (p<0.0001), VAFI and SOF = 0.25 (p<0.0001). The components of each frailty index are described in Figure 2.

Over a mean follow-up of 6±2 years, 17% (N=33,223) experienced all-cause mortality, 5% (N=10,115) CVD mortality, 1% (N=2,400) incident stroke, 2% (N=3,593) incident MI, and 9% (N=15,816) incident HF events. The associations between frailty and the primary outcomes of all-cause and CVD mortality are shown in Table 2. Using the MVP-FI, for CVD mortality, the respective HRs for pre-frail and frail were 1.76 (1.65-1.88) and 3.65 (3.43-3.90). For all-cause mortality, the multivariable-adjusted hazard ratio (HR, 95% CI) relative to robust individuals, was 1.66 (1.61-1.72) for pre-frail and 3.05 (2.95-3.16) for frail individuals. Findings were similar for pre-frail and frail individuals using the VA-FI and SOF indices (Table 2).

**Table 2.** Cox proportional hazard models for the association between frailty group and mortality.

| Outcome | Frailty Definition |  | No. Events | Total No. | HR (95% CI)* | P-value | c** |
| --- | --- | --- | --- | --- | --- | --- | --- |
| All-cause mortality | MVP-FI | Robust | 4,717 | 55,487 | 1.00 (Referent) | --- | 0.739 |
|  |  | Pre-frail | 13,060 | 79,835 | 1.66 (1.61-1.72) | <0.0001 |  |
|  |  | Frail | 15,446 | 55,366 | 3.05 (2.95-3.16) | <0.0001 |  |
|  | VA-FI | Robust | 7,196 | 78,004 | 1.00 (Referent) | --- | 0.741 |
|  |  | Pre-frail | 10,931 | 64,628 | 1.59 (1.54-1.64) | <0.0001 |  |
|  |  | Frail | 15,096 | 48,056 | 2.94 (2.85-3.04) | <0.0001 |  |
|  | SOF Index | Robust | 13,574 | 98,434 | 1.00 (Referent) | --- | 0.730 |
|  |  | Pre-frail | 13,538 | 70,167 | 1.57 (1.54-1.61) | <0.0001 |  |
|  |  | Frail | 5,154 | 18,091 | 2.60 (2.52-2.69) | <0.0001 |  |
| CVD death | MVP-FI | Robust | 1,217 | 55,487 | 1.00 (Referent) | --- | 0.779 |
|  |  | Pre-frail | 3,775 | 79,835 | 1.76 (1.65-1.88) | <0.0001 |  |
|  |  | Frail | 5,123 | 55,366 | 3.65 (3.43-3.90) | <0.0001 |  |
|  | VA-FI | Robust | 1,793 | 78,004 | 1.00 (Referent) | --- | 0.781 |
|  |  | Pre-frail | 3,162 | 64,628 | 1.70 (1.60-1.81) | <0.0001 |  |
|  |  | Frail | 5,160 | 48,056 | 3.52 (3.33-3.73) | <0.0001 |  |
|  | SOF Index | Robust | 4,142 | 98,434 | 1.00 (Referent) | --- | 0.762 |
|  |  | Pre-frail | 4,094 | 70,167 | 1.58 (1.51-1.65) | <0.0001 |  |
|  |  | Frail | 1,569 | 18,091 | 2.63 (2.48-2.79) | <0.0001 |  |

Abbreviations: SOF, Study of Osteoporotic Fractures FI, frailty index; CVD, cardiovascular disease.
\*Adjusted for age, sex, smoking status, statin use, antihypertensive treatment, history of hyperlipidemia, days between MVP baseline and lifestyle surveys, and MVP baseline survey year.
\*\* Harrell's c-statistic

The associations between frailty and secondary outcomes of incident stroke, MI, and HF are shown in Table 3. The results demonstrate statistically significant associations for the MVP-FI, VA-FI, and SOF with incident CVD events with a dose-response relationship of greater frailty leading to a greater hazard for events for each frailty measure. For example, in multivariable-adjusted models, HR (95%CI) for incident stroke were 1.51 (1.34-1.70) and 2.29 (2.03-2.58), for pre-frail and frail participants using the MVP-FI, with similar patterns for the VA-FI, and SOF.

**Table 3.** Cox proportional hazard models for the association between frailty group and CVD outcomes.

| Outcome | Frailty Definition |  | No. Events | Total No. | HR (95% CI)* | P-value | c** |
| --- | --- | --- | --- | --- | --- | --- | --- |
| MI | MVP-FI | Robust | 531 | 54,383 | 1.00 (Referent) | --- | 0.720 |
|  |  | Pre-frail | 1,482 | 75,182 | 1.69 (1.53-1.87) | <0.0001 |  |
|  |  | Frail | 1,580 | 47,869 | 2.67 (2.41-2.95) | <0.0001 |  |
|  | VA-FI | Robust | 976 | 76,487 | 1.00 (Referent) | --- | 0.716 |
|  |  | Pre-frail | 1,302 | 60,369 | 1.43 (1.31-1.56) | <0.0001 |  |
|  |  | Frail | 1,315 | 40,578 | 2.22 (2.03-2.43) | <0.0001 |  |
|  | SOF Index | Robust | 1,573 | 93,093 | 1.00 (Referent) | --- | 0.706 |
|  |  | Pre-frail | 1,483 | 64,619 | 1.29 (1.20-1.38) | <0.0001 |  |
|  |  | Frail | 446 | 16,043 | 1.54 (1.39-1.72) | <0.0001 |  |
| Stroke | MVP-FI | Robust | 405 | 54,720 | 1.00 (Referent) | --- | 0.710 |
|  |  | Pre-frail | 997 | 76,717 | 1.51 (1.34-1.70) | <0.0001 |  |
|  |  | Frail | 998 | 49,854 | 2.29 (2.03-2.58) | <0.0001 |  |
|  | VA-FI | Robust | 670 | 76,972 | 1.00 (Referent) | --- | 0.707 |
|  |  | Pre-frail | 893 | 61,788 | 1.45 (1.30-1.60) | <0.0001 |  |
|  |  | Frail | 837 | 42,531 | 2.06 (1.85-2.30) | <0.0001 |  |
|  | SOF Index | Robust | 1,052 | 94,817 | 1.00 (Referent) | --- | 0.701 |
|  |  | Pre-frail | 980 | 66,251 | 1.33 (1.22-1.45) | <0.0001 |  |
|  |  | Frail | 315 | 16,533 | 1.75 (1.54-1.99) | <0.0001 |  |
| CHF | MVP-FI | Robust | 2,344 | 54,218 | 1.00 (Referent) | --- | 0.710 |
|  |  | Pre-frail | 6,587 | 74,161 | 1.69 (1.61-1.77) | <0.0001 |  |
|  |  | Frail | 6,885 | 44,595 | 2.95 (2.81-3.09) | <0.0001 |  |
|  | VA-FI | Robust | 3,600 | 76,847 | 1.00 (Referent) | --- | 0.715 |
|  |  | Pre-frail | 5,968 | 60,156 | 1.72 (1.65-1.80) | <0.0001 |  |
|  |  | Frail | 6,248 | 35,971 | 3.07 (2.94-3.21) | <0.0001 |  |
|  | SOF Index | Robust | 6,958 | 92,322 | 1.00 (Referent) | --- | 0.692 |
|  |  | Pre-frail | 6,402 | 62,317 | 1.39 (1.34-1.44) | <0.0001 |  |
|  |  | Frail | 2,071 | 14,858 | 1.99 (1.89-2.09) | <0.0001 |  |

Abbreviations: CVD, cardiovascular disease; SOF, Study of Osteoporotic Fractures FI, frailty index; MI, myocardial infarction; CHF, congestive heart failure.
\*Adjusted for age, sex, smoking status, statin use, antihypertensive treatment, history of hyperlipidemia, days between MVP baseline and lifestyle surveys, and MVP baseline survey year.
\*\* Harrell's c-statistic

**Table 4.** Cox proportional hazard models for the association between frailty group and mortality, stratified by sex.

| Sex | Frailty Definition |  | Total No. | All-Cause Mortality |  |  |  | CVD Death |  |  |  |
| --- | --- | --- | --- | --- | --- | --- | --- | --- | --- | --- | --- |
|  |  |  |  | No. Events | HR (95% CI)* | P-value | c** | No. Events | HR (95% CI)* | P-value | c** |
| Female | MVP-FI | Robust | 3,330 | 102 | 1.00 (Referent) | --- | 0.790 | 21 | 1.00 (Referent) | --- | 0.853 |
|  |  | Pre-frail | 4,725 | 279 | 1.35 (1.07-1.7) | 0.01 |  | 61 | 1.27 (0.77-2.1) | 0.35 |  |
|  |  | Frail | 3,797 | 514 | 2.64 (2.11-3.3) | <.0001 |  | 144 | 3.03 (1.88-4.86) | <.0001 |  |
|  | VA-FI | Robust | 5,114 | 167 | 1.00 (Referent) | --- | 0.794 | 39 | 1.00 (Referent) | --- | 0.849 |
|  |  | Pre-frail | 3,887 | 262 | 1.46 (1.2-1.79) | 0.0002 |  | 58 | 1.16 (0.76-1.76) | 0.49 |  |
|  |  | Frail | 2,851 | 466 | 2.79 (2.3-3.39) | <.0001 |  | 129 | 2.5 (1.69-3.68) | <.0001 |  |
|  | SOF Index | Robust | 5,346 | 312 | 1.00 (Referent) | --- | 0.783 | 84 | 1.00 (Referent) | --- | 0.840 |
|  |  | Pre-frail | 4,878 | 399 | 1.49 (1.29-1.74) | <.0001 |  | 97 | 1.39 (1.04-1.87) | 0.03 |  |
|  |  | Frail | 1,409 | 163 | 2.34 (1.93-2.83) | <.0001 |  | 40 | 2.29 (1.56-3.37) | <.0001 |  |
| Male | MVP-FI | Robust | 52,157 | 4,615 | 1.00 (Referent) | --- | 0.732 | 1,196 | 1.00 (Referent) | --- | 0.772 |
|  |  | Pre-frail | 75,110 | 12,781 | 1.67 (1.61-1.73) | <.0001 |  | 3,714 | 1.77 (1.66-1.89) | <.0001 |  |
|  |  | Frail | 51,569 | 14,932 | 3.06 (2.96-3.17) | <.0001 |  | 4,979 | 3.67 (3.43-3.91) | <.0001 |  |
|  | VA-FI | Robust | 72,890 | 7,029 | 1.00 (Referent) | --- | 0.735 | 1,754 | 1.00 (Referent) | --- | 0.774 |
|  |  | Pre-frail | 60,741 | 10,669 | 1.59 (1.54-1.64) | <.0001 |  | 3,104 | 1.72 (1.62-1.82) | <.0001 |  |
|  |  | Frail | 45,205 | 14,630 | 2.95 (2.86-3.04) | <.0001 |  | 5,031 | 3.55 (3.35-3.76) | <.0001 |  |
|  | SOF Index | Robust | 93,088 | 13,262 | 1.00 (Referent) | --- | 0.723 | 4,058 | 1.00 (Referent) | --- | 0.754 |
|  |  | Pre-frail | 65,289 | 13,139 | 1.58 (1.54-1.62) | <.0001 |  | 3,997 | 1.58 (1.52-1.65) | <.0001 |  |
|  |  | Frail | 16,682 | 4,991 | 2.61 (2.53-2.7) | <.0001 |  | 1,529 | 2.64 (2.49-2.8) | <.0001 |  |

Abbreviations: SOF, Study of Osteoporotic Fractures FI, frailty index; CVD, cardiovascular disease.
\*Adjusted for age, smoking status, statin use, antihypertensive treatment, history of hyperlipidemia, days between MVP baseline and lifestyle surveys, and MVP baseline survey year.
\*\* Harrell's c-statistic

Figure 1 demonstrates the association of MVP-FI frailty group with all-cause and CVD mortality stratified by age and sex. A nearly 3-fold increase in risk at 50-59 years (HR 2.73; 95%CI 2.36-3.15) was seen in frail participants. The highest risk was among those aged 60-69 years (HR 3.25; 95%CI 3.05-3.46) with a subsequent graded decline by advancing decade. Similar patterns were seen for CVD mortality. When stratified by sex, the hazard of all-cause mortality and CVD mortality was slightly higher in men compared to women for both the pre-frail and frail groups. These patterns remained consistent across frailty indices with some attenuation where sample size was limited.

**Figure 1.**
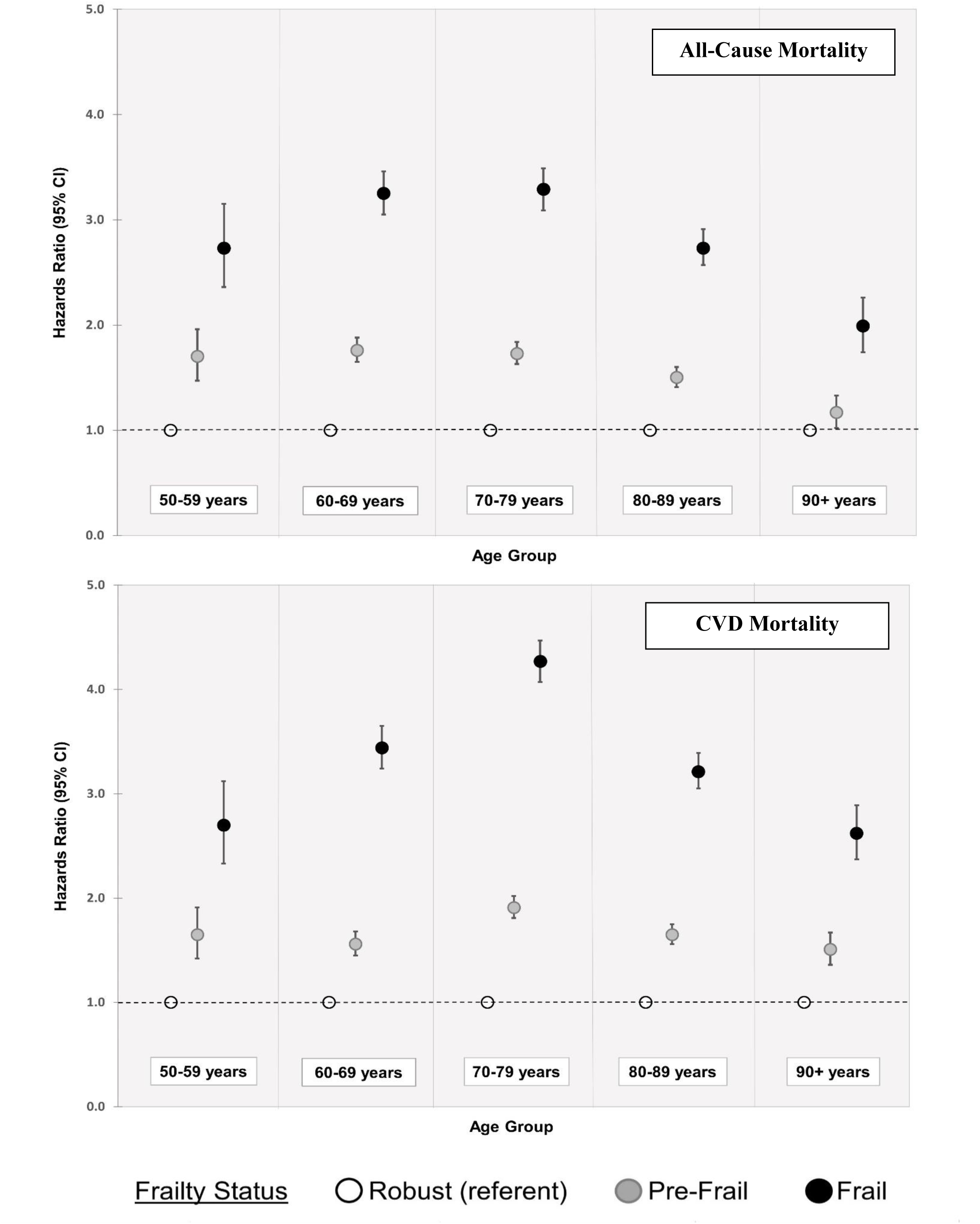
Hazards ratios (HR) for the association between frailty group and *all-cause mortality* (upper panel) and *CVD mortality* (lower panel), stratified by 10-year age group.

**Figure 2.**
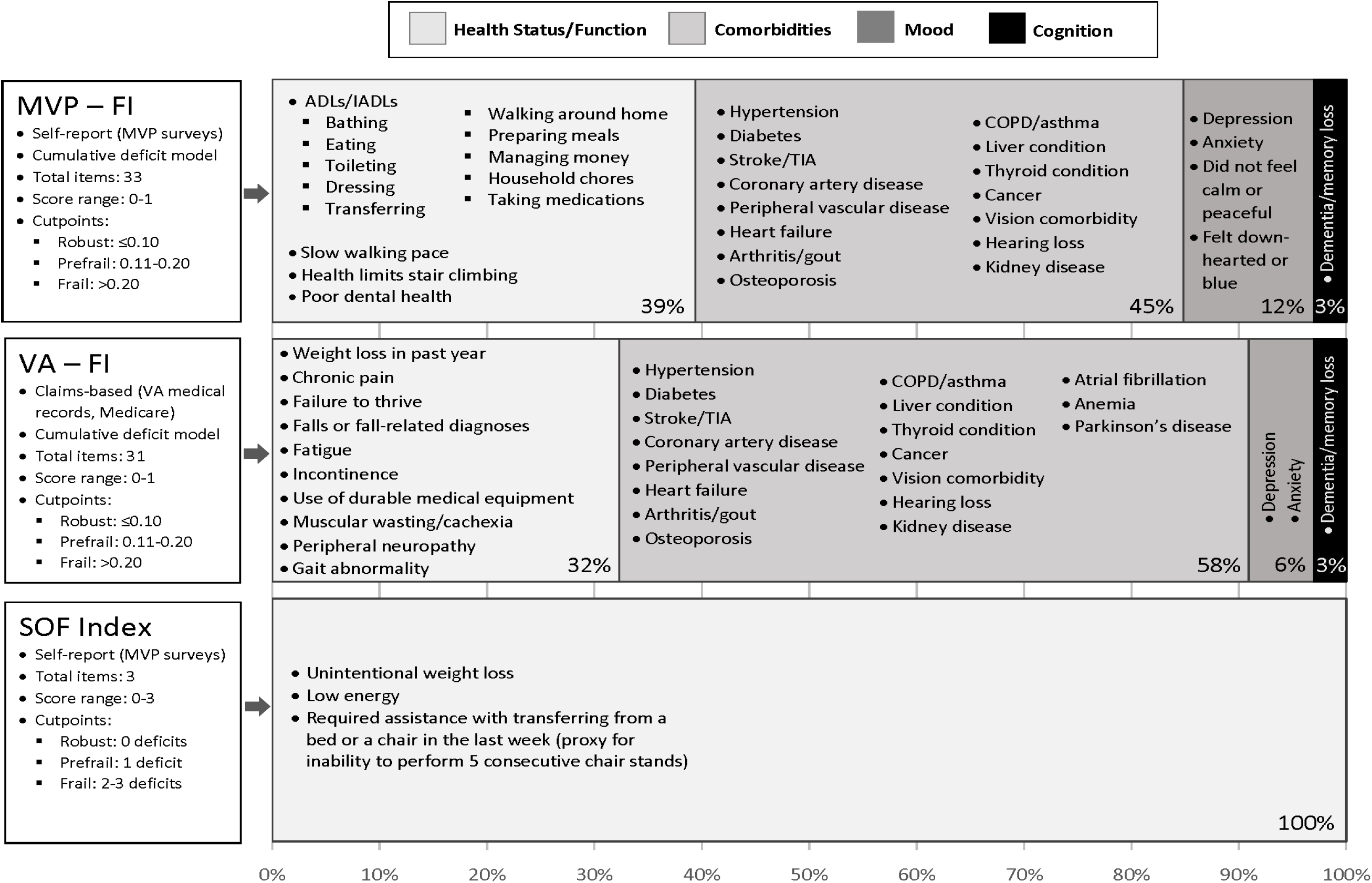
Graphical Overview of frailty indices (MVP-FI, VA-FI, and SOF Index). Percentages shown indicate the proportion of total deficits for each frailty index attributed to each domain (health status/function, comorbidities, mood, and cognition).

## DISCUSSION

In a contemporary, deeply-phenotyped cohort of US veterans, we demonstrate that both pre-frailty and frailty are significantly associated with all-cause and CVD mortality using three distinct measures of frailty. This was true despite the use of differing parameters extracted from self-reported and claims-based data for the MVP-FI, VA-FI, and SOF tools. A similar pattern was seen for secondary endpoints such as stroke, MI, and HF. We also identified a graded increase in risk of adverse events with increasing frailty status.

### In the Context of Current Literature

Our data demonstrated the burden of frailty was consistent with prior community dwelling populations.^26^ Prior literature has also demonstrated a significant association between frailty and all-cause mortality. Moreover, a higher number of frailty markers or increasing frailty severity results in a dose-response increase in risk of all-cause mortality across different populations.^27–30^ The risk of mortality associated with frailty in prevalent CVD and the incremental association between frailty and CVD mortality have been previously described.^31–34^ In a meta-analysis of patients (N=154,696) with prevalent CVD or at high risk of CVD enrolled in 14 randomized clinical trials, Farooqi et al demonstrated that frailty assessed by the cumulative deficit approach was associated with increased CVD mortality risk after adjusting for baseline comorbidities (HR 2.06, 95% CI 1.76-2.42).^35^ Veronese et al reported a similar relationship between frailty and CVD mortality in a meta-analysis of 18 cohorts with a nearly four-fold increase in risk of CVD mortality among those participants identified as frail (HR 3.89; 95% CI 2.40-6.34).^36^ Prior data from VA has demonstrated that among US Veterans aged ≥65 years, the VA-FI was predictive of increased risk of all-cause and CVD mortality.^37^ The present analysis uniquely extends the current literature by demonstrating the consistent prognostic utility of frailty regardless of how it is measured: either deficit accumulation or physical phenotype, using claims data or self-report.

Stratification by age identified that frailty in younger age individuals (aged 50-59), typically not considered for frailty assessments, may portend even greater risk than in older adults. The prevalence of frailty has been described as ranging from 3.9% to 63% among individuals aged 18-65.^38^ In our data, 27% of those aged 50-59 were frail using the MVP-FI, and the hazard of dying from a CVD event was nearly three-fold. These findings lay the groundwork for advancing the concept of frailty as a global measure of physiologic age beyond the reliance on numeric age alone, and its potential utility in younger patients for improving CVD risk stratification.

Limited data are available on the relationship between of frailty and incident CVD.^12,16^ Sergi et al showed a 25% higher risk of CVD events comparing pre-frail to non-frail individuals in a population of 1,567 participants aged 65-96 years (HR 1.25; 95% CI 1.05-1.64). Pre-frail resulted in a nearly 80% increase in risk of CVD compared to those who were pre-frail or robust.^39^ Damluji et al reported a similar association between frailty and CVD events among 3,259 participants of the National Health and Aging Trends Study. Specifically, acute MI (HR 1.95, 95% CI 1.31-2.90) and stroke (HR 1.71, 95% CI 1.34-2.17), risk was nearly two-fold higher in frail participants.^40^ Both of these studies demonstrated these associations using physical frailty indices. Our study extends these data in a large, contemporary cohort using both deficit accumulation and physical frailty tools that provide similar prognostic information regarding incident CVD risk.

Notably, in addition to considering outcomes such as mortality and atherosclerotic CVD, we expand the literature with our investigation of the association of frailty with HF. Frailty is known to be highly prevalent in patients with HF (36.2-52.8%).^16^ Irrespective of type, outcomes are worse among those with prevalent HF who are also frail.^41–44^ Prior to the current study, there has been paucity of literature frailty as a prognostic risk factor for incident HF.

### Potential Mechanisms

CVD as a risk factor for frailty and frailty as a predictor of CVD and mortality has been attributed to the bidirectional association between frailty and CVD.^16^ The overlapping mechanisms that underlie both frailty and CVD, including inflammation, insulin resistance, and cellular senescence have been described as key players in the development and progression of both CVD and frailty.^12^ Systemic inflammation is a shared pathophysiologic mechanism leading to frailty, changes in muscle physiology with aging, and subclinical CVD impacting multiple organ systems.^12^ This leads to a cyclical relationship of worsening functional status which then leads to progression of CVD risk factors such as adiposity, metabolic syndrome, and chronic low-grade inflammation.^12,16^

### Clinical Implications and Future Directions

Our data support the use of frailty as a valuable tool for assessing the cardiovascular health of older adults using the most readily available tool. This may be a detailed questionnaire, EHR claims, or self-reported function, and can be tailored to the needs and resources of a particular clinic or health system. For example, in the UK,^45^ frailty has already been included as an automated tool in the electronic health record across all of primary care. Similar strategies can be taken in other healthcare systems to readily identify patients at risk who will benefit from targeted preventive therapies.

The similar performance of different frailty measures as shown in our study is important: different measures will identify different individuals as frail, particularly when comparing physical and deficit accumulation models. This can be a challenge to clinicians given the array of different frailty instruments that are available as well as limited time to undertake an assessment. However, regardless of the variations in frailty assessment tools, the identification of frailty itself portends greater risk of mortality and CVD events. Deficit accumulation frailty indices using EHR data, such as the VA-FI, present one means of addressing time limitations in frailty assessment. Frailty assessment can then follow up an automated assessment with an in-person evaluation. It is worth noting that frailty tools based on different theories (i.e., physical phenotype and deficit accumulation) by design have poor concordance and a combination of frailty assessment tools may be needed to thoroughly screen patients; however, utilization of any tool(s) at a clinician’s disposal can sufficiently risk stratify patients. Supplemental Figure 2 shows a proposed clinic workflow to implement different types of frailty assessment and maximize new information on individuals whose frailty and associated health risks may be missed.

Finally, our findings were robust across age groups. While frailty assessment is often considered a beneficial tool for those aged ≥65, and particularly those ≥80 years—exceeding maximum age in most CVD risk calculators—we found that frailty assessment in middle-aged adults may be of clinical utility in risk stratification as well. Further prospective studies are needed to support these findings.

### Strengths and Limitations

Our study benefits from a large sample of volunteer participants with extensive follow-up both through a large, integrated health organization linked to the Centers for Medicare and Medicaid Services (CMS). However, these CMS data were available in the older subgroups of our study which may have resulted in underestimation of frailty level in younger groups using the VA-FI.

Our frailty assessment was limited to items covered in survey instruments, which did not include direct physical measures of strength or gait speed nor common blood-based laboratory tests. The sample included was also predominantly male and white, which may limit generalizability.

## CONCLUSION

Frailty, irrespective of how it was measured, is predictive of mortality and CVD, even among those aged 50-59. These findings suggest that clinicians and researchers should use the tool that is most convenient to incorporate frailty into practice.

## Data Availability

Data are available to approved investigators per VA IRB rules and regulations.

## ABBREVIATIONS

(CVD): Cardiovascular disease
(HF): Heart Failure
(MI): Myocardial infarction
(MVP): Million Veteran Program
(FI): Frailty index
(VA): Veterans Administration
(CMS): Centers for Medicare and Medicaid Services

## CLINICAL PERSPECTIVE

- Among Veterans 50 years and older, frailty was associated with a higher risk of CVD and all-cause mortality and adverse events such as incident myocardial infarction, stroke, and heart failure. This association was seen irrespective of type of frailty assessment tool used (i.e., physical phenotype versus deficit accumulation).
- Frailty is a valuable tool for assessing the cardiovascular health of older adults. Among the varying frailty assessment tools such as detailed questionnaire, EHR claims, or self-reported function, the most readily available one can be used based on the needs and resources of a particular clinic or health system.
- Assessment of frailty to evaluate CVD risk may also be useful in younger adults.

## ACKNOWLEDGEMENTS

We thank Timothy Treu, MPH for his assistance with the data quality check process.

Support for VA/CMS data provided by the Department of Veterans Affairs, VA Health Services Research and Development Service, VA Information Resource Center (Project Numbers SDR 02-237 and 98-004).

Veterans Health Administration, Center of Excellence for Suicide Prevention. Joint Department of Veterans Affairs and Department of Defense Mortality Data Repository. Data compiled from the National Death Index. https://www.mirecc.va.gov/suicideprevention/documents/VA_DoD-MDR_Flyer.pdf; Extract November 10, 2021.

## SUPPLEMENTAL MATERIAL

### Figures

Supplemental Figure 1. Study Sample Selection.

Supplemental Figure 2. Proposed clinic workflow to integrate deficit accumulation and physical frailty assessment.

### Tables

Supplemental Table 1. Summary of MVP Baseline and Lifestyle Questionnaire Items Used to Create the MVP Frailty Index.

Supplemental Table 2. Summary of MVP Baseline and Lifestyle Questionnaire Items Used to Create the SOF Index.

Supplemental Table 3. Cox proportional hazard model results for the association between frailty group and mortality, stratified by age group.

Supplemental Table 4. Cox proportional hazard model results for the association between frailty group and mortality among participants who completed their Baseline and Lifestyle Surveys within a 2-year period.

